# Single-cell RNA analysis reveals the cell atlas of human intracranial aneurysm and rupture-related inflammation features

**DOI:** 10.1101/2023.05.03.23289486

**Authors:** Hang Ji, Yue Li, Haogeng Sun, Ruiqi Chen, Ran Zhou, Anqi Xiao, Yongbo Yang, Rong Wang, Chao You, Yi Liu

**Affiliations:** Department of Neurosurgery, West China Hospital, Sichuan University, Chengdu, 610041, China; Department of Neurosurgery, State Key Laboratory of Biotherapy and Cancer Center, West China Hospital, Sichuan University, Chengdu, 610041, China; Department of Neurosurgery, Nanjing Drum Tower Hospital, The Affiliated Hospital of Nanjing University Medical School, Nanjing, 210008, China; Department of Neurosurgery, Beijing Tiantan Hospital, Capital Medical University, Beijing, 100070, China

**Keywords:** Human intracranial aneurysm, single-cell transcriptome, pericyte, inflammation, macrophage

## Abstract

**Background:** Intracranial aneurysms (IA) is a common condition and may ultimately result in life-threatening hemorrhagic strokes. A precise understanding of the cellular and gene expression perturbations in human IA tissue may enlighten additional therapeutics for unruptured IA.

**Methods:** A total of 21,332 qualified cells were obtained from four cell-sparse ruptured and unruptured human IA tissues. Detailed cell atlas and dynamics, gene expression perturbations, and inflammation features were thoroughly investigated using multiple machine learning-based algorithms.

**Results:** Endothelial cells, smooth muscle cells (SMCs), fibroblasts and, for the first time, pericytes have been identified in human IA tissue. A significant proportion of immune cells are also identified, with the number of monocyte/macrophages and neutrophils being notably higher in ruptured IA. By leveraging external datasets, macrophages characterized by transcriptional activation of NF-κB and HIVEP2 is most strongly associated with IA rupture. Interestingly, the recruitment and activation of macrophages and their functional characteristics in terms of TNFα and chemokine production remain consistent between unruptured and ruptured IA.

**Conclusions:** This study provides insights into the pathophysiology and molecular underpinnings of the IA wall and may motivate novel therapeutic options for unruptured IA.

## Introduction

Intracranial aneurysm (IA) represents a group of diseases manifested as confined pathological outpouchings of the Circle of Willis, with an incidence of approximately 3∼5% in the general population(1-3). Spontaneous subarachnoid hemorrhage (SAH) caused by IA rupture is a highly life-threatening condition that often affects middle-aged people, with mortality or disability occurring in over 50% of patients(4-6). Worryingly, an increasing number of people are found to carry single or multiple unruptured IAs during routine screening with the spread of angiography(7, 8). Despite recent advances in the clinical management of IA, endovascular and surgical aneurysm repair, or merely conservative management have inherent risks of complications. To facilitate the treatment of IA, it is essential to have a thorough understanding of its pathological mechanism.

Recent advances have yielded important insights into the pathophysiological alterations of IA. For instance, histological studies based on patients’ and murine model-derived IA tissues highlight decellularization, internal elastic lamina remodeling, and chronic inflammation during the dilation of vascular wall(7, 9-11). Magnetic resonance imaging-based studies suggest an association between circumferential aneurysm wall enhancement with inflammation and macrophage infiltration, which serves as a sign of increased risk of rupture(7, 12, 13). Over the last decade, computational-based hemodynamics studies have elucidated the hemodynamic perturbations associated with the onset and progression of IA, adding critical evidence for the morphological and functional conversions of endothelial and SMC induced by wall shear stress(9, 14, 15). Recently, genome-wide association studies and other high-throughput screening-based studies have illuminated genetic risk locoes, genes, and modifiable epigenetic factors associated with IA pathology(6, 16-19). Overall, these scientific advances have provided a comprehensive framework for understanding the pathological mechanisms of IA.

Etiologically, the controversy remains as to whether IA is an acquired lesion caused by local hemodynamic stress or a manifestation of inherent vulnerability in the vessel wall(1, 3, 6, 18). In any event, the type and state of cells within the intima and media layers of the IA wall exert a profound impact on disease progression(7). Single-cell transcriptome sequencing has partially mitigated the cellular sparsity in studying the cerebrovasculature, enabling unprecedentedly detailed profiles of cellular and gene expression under both normal and pathological conditions(20, 21). Recent studies utilizing murine and human IA tissue have pinpointed the role of macrophages, which are also responsible for bleeding in intracranial arteriovenous malformations and thoracic aortic aneurysms(20, 22-24). Nevertheless, the cellular vulnerabilities of general human IA tissue and inflammation dysregulations associated with IA rupture remain largely unknown. In this perspective, we conducted single-cell transcriptome sequencing on patient-derived IA samples at different pathological stages to investigate the fundamental molecular and cellular mechanisms and inflammation features implicated in IA rupture.

## Materials and methods

### IA sample and external data collection

This study was approved by the Ethics Review Committee of West China Hospital, Sichuan University, and was performed in accordance with Good Clinical Practice guidelines and the 1964 Helsinki Declaration and its later amendments. All participants were informed and signed a written consent form priorly. All data used were de-identified. Patients were diagnosed as IA angiographically before bypass surgery or clipping. Fresh IA tissues were obtained from 3 SAH and 1 unruptured IA patients. Tissues were obtained in close collaboration with two senior neurosurgeons with extensive experience in IA surgery to minimize destruction. Thrombus-containing IA tissue was avoided. The obtained IA tissue was immediately flushed of erythrocytes. Patients were excluded if they had a heritable arterial disease (Marfan syndrome, Ebler-Danlos syndrome, pseudoxanthoma elasticum, autosomal dominant polycystic kidney disease, neurofibromatosis type I). Two external microarray-based IA datasets were retrieved from the Gene Expression Omnibus, including GSE13353 and GSE122897(25, 26). Positive and negative hits of IL-4- and IFNγ-polarized macrophage, and gene signatures of different conditions-polarized macrophages were retrieved from GSE47189(27).

### Single-cell RNA sequencing sample preparation

Fresh specimens were placed in chilled MACS tissue storage solution (Miltenyi, Germany) and transported to the laboratory on ice. The IA tissue was slightly flushed with prechilled DMEM (Sigma-Aldrich, USA) to remove erythrocytes. Then, the tissue was fragmented and digested in mixed digestion buffer (normal Dulbecco’s modified Eagle’s medium containing 15 mg/ml collagenase type II + 500 ug/ml elastase) for approximately 30 mins at room temperature. Tissue suspension was filtered through a sterilized a 70-μm followed by a 40-μm cell strainer to isolate debris. Isolated cells were collected by centrifugation at 500g for 5 minutes at 4°C, and the supernatant was carefully aspirated. The cell pellet was resuspended in Gibco ACK Lysis Buffer (Thermo Fisher Scientific, USA) for 3 minutes to lyse residual erythrocytes. Cells were then collected by centrifugation at 500g for 5 minutes and washed three times with sterile RNase-free phosphate-buffered saline (PBS) (Beyotime, China) containing 0.04% BSA (Beyotime, China). Thereafter, cells were pelleted and resuspended in PBS containing BSA. Cell viability and concentration were assessed automatically using the Countstar Rigel S2 system (Countstar China). The microfluidics-based cDNA library construction and quality control, and sequencing based on the Novaseq6000 platform were conducted by Biomarker Technologies CO., Ltd. (Beijing, China).

### Single-cell RNA sequencing data analysis

Count matrices of cell-by-gene was created using Cellranger (v7.1.0). Raw sequencing reads were aligned to the pre-mRNA annotated by homo sapiens reference genome GRCh38. Seurat framework was employed to manage the downstream analysis. DoubletFinder was employed for potential doublet detection at a criteria of 7.5% doublet formation rate based on the recommendation of 10× Genomics(28). A minimum of 500 genes and 15% mitochondrial cutoff were used to remove low quality cells. The SCTransform workflow was used for count normalization. Diagonalized canonical correlation analysis (CCA) and mutual nearest neighbors (MNN) were performed for batch correction(29). The scaled data with variable genes identified by Seurat were used to perform principal component analysis (PCA). The first 30 principal components of the PCA were used for clustering and cell identification. Annotation of cell clusters leveraged the automatic approach, Azimuth(30), followed by manual annotation using conserved cell type markers. Pseudotime analysis was performed using the R package ‘monocle3’(31). CellChat and cellphonedb (V3.0) were employed to predict cell-cell communication(32, 33). SCENIC was used to identify activated transcription factors(34). Weighted gene co-expression network analysis (WGCNA) was performed to identify the intrinsic gene expression program(35). As the single-cell count matrix is sparse, pseudocells was constructed as the averages of 5-10 randomly chosen cell within each cluster. Functional enrichment analysis was performed using the webtool Metascape(36) (https://www.metascape.org/).

### Correlation between immune cells with IA rupture

Marker genes of each type of cell were defined as those average log2FC > 0.5, Pct.2 < 0.15, and adjusted p-values < 0.1. Single-sample gene set enrichment analysis (ssGSEA) were employed to estimate the abundance of each type of cell in bulk sample. Receiver operating characteristic curves (ROC) and corresponding area under the curves (AUC) were calculated for estimating the correlation.

### Statistics

All statistics were performed using the R software (v4.2.1). Mann-Whitney U test was employed for the comparison of non-normally distributed variables. The prediction accuracy was estimated using ROC and corresponding AUC. A two-sided P-value <= 0.05 was defined as statistically significant unless otherwise specified.

## Results

### Cellular and molecular profiles of IA

The raw data from each of the four samples were subjected to low quality cell filtering and noise correction, and then co-embedded (29, 37). A total of 21,332 qualified cells were obtained (Supplementary Figure 1A-C). Unbiased cell clustering analysis was performed to identify 21 cell clusters (Supplementary Figure 1D-F). Based on established cell identity specific transcriptomic biomarkers, 8 major cell types were first assigned (Figure 1A, B). Vascular cells including endothelial cell (*CLDN5, PECAM1*)(21), pericyte (*ABCC9, KCNJ8*)(20), fibroblast (*DCN*)(20) and SMC (*MYH11, CNN1*)(23), accounted for approximately 4.8% of the total cells. Immune cells made up the majority of the IA tissue, including granulocyte, mostly neutrophil (*S100A8, CXCL8*)(22, 38), monocyte/macrophage/dendritic cell (*CD14, CD16*)(39), T cell (*CD3E, CD3G*)(21), NK (*KLRF1, GZMA*)(21), B cell (*CD79A, MS4A1*)(21) and mast cell (*TPSAB1, CPA3*)(21). No type of cell was absent from a sample, except for fibroblasts, and their proportions in rupture (IA1∼3) and unruptured (IA4) tissues varied considerably (Figure 1C).

**Figure 1.**
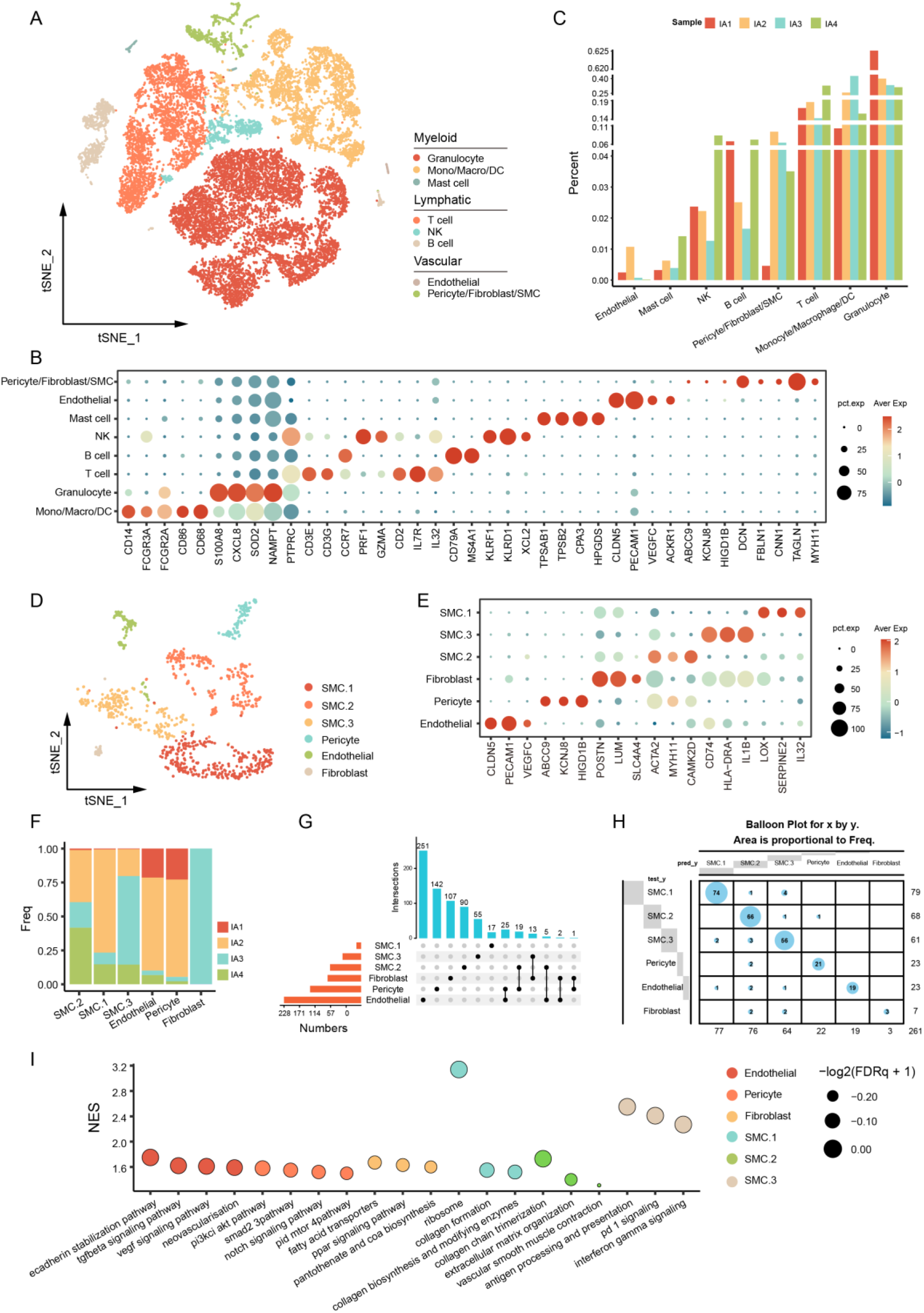
Cells and gene expression features of human IA tissue. (**A**) tSNE visualization of major cell types. (**B**) Dot plot exhibiting conserved cell identity biomarkers. (**C**) Bar plot exhibiting the proportion of different cells in each sample. (**D, E**) Vascular cell types and states, and their feature genes. (**F**) Proportion of vascular cells in each sample. (**G**) The intersection of feature genes (log2FC > 1) of each cell type. (**H**) Random forest-based cell classifier for the identification of major cell types. (**I**) GSEA analysis of pathways enriched in each type of cell.

### Vascular cells in IA tissue

Normal cerebral arteries and arterioles are composed of endothelial cell, SMC, and perivascular fibroblast(20, 21). Given that IA was characterized by pathological dilation of vascular wall and decellularization, the presence and status of these cells remain unclear(40). To gain further insights, the vascular cells identified in the initial clustering were re-clustered at a higher resolution, with 4 types of vascular cells including endothelial cell (n = 89), pericyte (n = 92), fibroblast (n = 26), and smooth muscle cell (SMC, n = 826) being identified (Figure 1D).

Endothelial cells form the blood-facing surface of the intracranial artery, and is stressed and injured by the wall shear stress(41). In this scenario, endothelial cells were marked by increased expression of *CLDN5, CD31*(*PECAM1*), and a recently identified endothelial marker gene, *VEGFC*(20) (Figure 1E). Genes up-regulated in endothelial were involved in the vascular development, tissue morphogenesis, cell migration and adhesion (Figure 1I, Supplementary Figure 2A).

Pericyte exists around microvessels and play a role in regulating blood flow and angiogenesis, and has been detected in the cerebral vasculature(20, 21, 42). We for the first time reported the presence of pericyte in human IA tissue. Pericyte was marked by *ABCC9* and *KCNJ8* and uniquely expressed *HIGD1B* (Pct.1 = 79.5%, Pct.2 = 1.9%, log_2_FC = 1.63), in line with previous findings(20). The identification of pericyte was based on two additional clues: 1. feature genes of pericytes had little overlap with other cells (Figure 1G), and 2. the random forest-based cell classifier suggested that this type of cell was different from others (Figure 1H). Pericytes had activated signaling pathways including PI3K-Akt-mTOR, SMAD2, and NOTCH signaling (Figure 1E, Supplementary Figure 2), which are essential for cell survival and differentiation(43). Previous study reported the presence of vasa vasorum in IA tissue(44), cell-cell communication analysis found the interaction between endothelial and pericyte through PDGFB-PDGFRB and TGFB1-TGFBR2 pathways, and predicted the involvement of TGFB3 in the endothelial TGFB signaling activation (Supplementary Figure 2C-E), corroborating the role of pericytes in neovascularization(42, 45).

IAs are characterized by loss of SMCs(10). We found that SMC cells can be further divided into three subgroups. SMC.1 up-regulated genes involved in extracellular matrix portraying (*ECM*) and inflammation (*LOX, IL32*, and *SERPINE2*), and had increased activity of signaling pathways associated with collagen biosynthesis and modifying (Figure 1E, I, Supplementary Figure 2F). SMC.2 highly expressed genes encoding actin (*ACTA2*) and classical SMC transcription factor (*MYH11*), indicating the retention of contraction function. Meanwhile, SMC.2 participated in the ECM organization via collagen production and organization (Figure 1E, I, Supplementary Figure 2G). SMC.3 was characterized by inflammation-associated genes (*CD74, HLA-DRA*, and *IL1B*), and the activation of signaling pathways including antigen presentation, interferon-gamma, and PD1 signaling (Figure 1E, I, Supplementary Figure 2H). Consistently, three types of SMCs were identified by monocle, including SMC contraction (*PDE3A*), SMC synthetic 1 and 2 (*TMSB10*), and SMC inflammation (*CD74, IL1B*) (Supplementary Figure 2I).

Perivascular fibroblasts play a role in maintaining the vascular integrity(46). Here, fibroblasts were all from the ruptured IA3 (Figure 1F), indicating that fibroblasts were scarce in IA tissue. Fibroblasts highly expressed *POSTN, SLC4A4* and *LUM* and enriched in pathways including fatty acid metabolism (Figure 1E, I, Supplementary Figure 2), indicating that they were involved in ECM remodeling and inflammation.

### Immune cells in IA tissue

Awful amounts of immune cells were found in the IA tissue(10, 47). These cells were re-clustered at a higher resolution, resulting in 18 clusters (Figure 2A, B). Myeloid cells including neutrophils and monocyte/macrophages/DC made up a significant proportion (43.2% and 20.1%) and lymphoid cells including NK cell, T cell, B cell and subclusters to a less extend (31.2%) (Figure 2B, C). Each type of immune cell was detected in both ruptured and unruptured IA tissue (Figure 2A, C).

**Figure 2.**
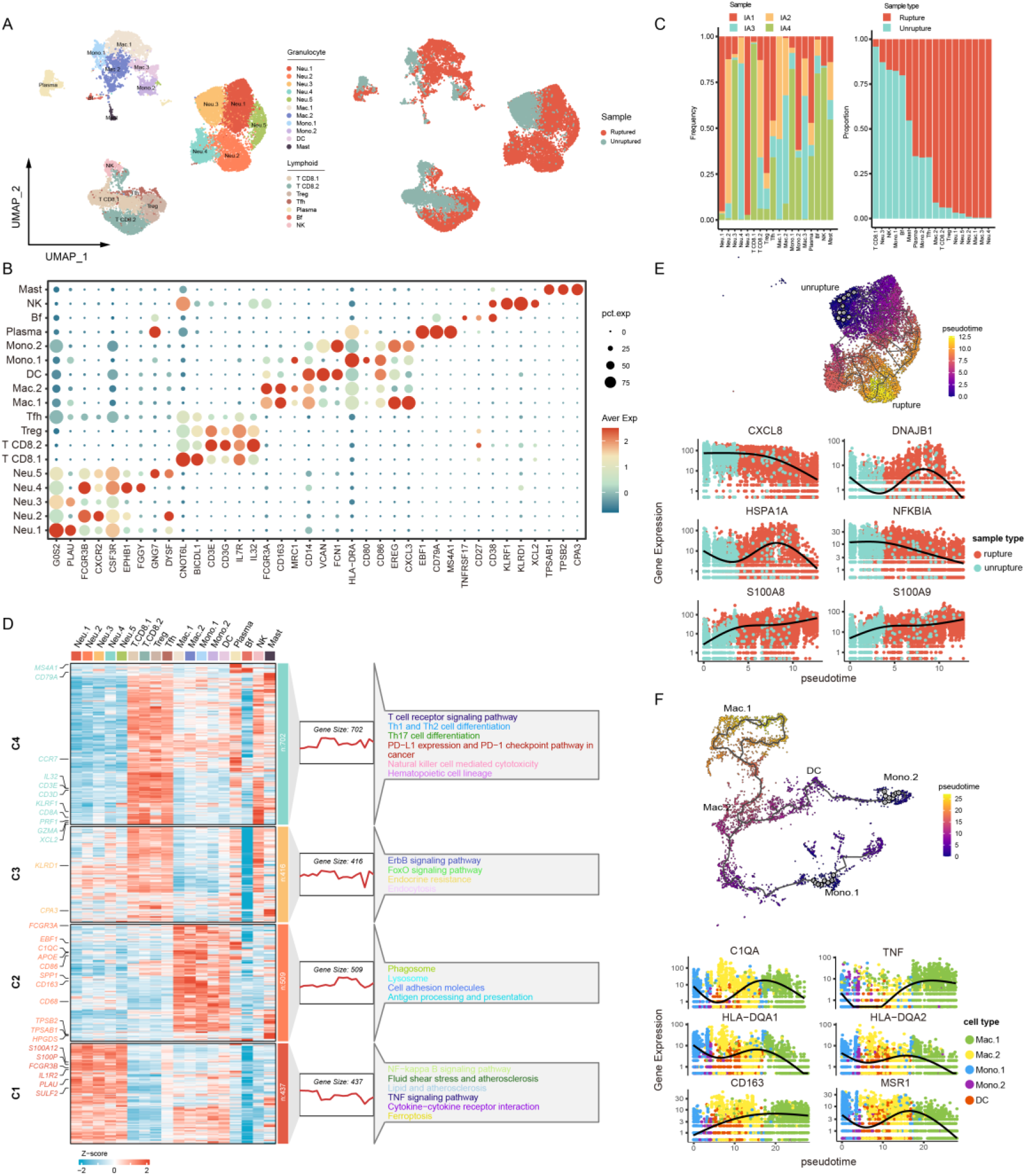
Immune cells and gene expression features. (**A**) UMAP visualization of immune cells and their distribution in ruptured (red) and unruptured (green) IAs. (**B**) The expression of conserved cell identity biomarkers. (**C**) Proportions of immune cells in each sample (left panel) and ruptured/unruptured IA tissue (right panel). (**D**) Gene expression patterns, feature genes, and enriched signaling pathways in major immune cell types. (**E**) Alterations in neutrophil gene expression in ruptured and unruptured samples. The start of the trajectories is artificially set as cells in unruptured sample. (**F**) Alterations in gene expression in different types of monocyte/macrophages. The start of the trajectories is set as Mono.1 and Mono.2.

### Granulocytes in the IA tissue

Granulocyte, namely neutrophil (*G0S2, PULA, FCGR3B*) in this condition(48, 49), was the most abundant immune cell in IA. Neutrophils was present even in the unruptured sample (31.1%). Genes highly expressed in neutrophils (Neu) were enriched in signaling pathways including NF-κB, TNF, and ferroptosis signaling (Figure 2D). Neutrophils are ferroptosis sensitive(50) and expressed higher *PTGS2* in contrast to other immune cells (Supplementary Figure 3A). Neu can be further divided into 5 subtypes (Figure 2A, B). Neu.1 was marked by increased expression of *G0S2* and *PLAU*, and was active in chemotaxis, migration and inflammatory response (Figure 2B, Supplementary Figure 3B). Neu.2 highly expressed *FCGR3B* and *CXCR2*, and had enriched pathways associated with migrating and serine/threonine activity (Figure 2B, Supplementary Figure 3B). Neu.3 highly expressed eicosanoid metabolizing enzyme encoding gene *ALOX5AP* (log2FC = 1.65, Pct.1 = 0.94, Pct.2 = 0.53) and *CXCL8* (log2FC = 1.55, Pct.1 = 1, Pct.2 = 0.73) (Figure 2B, Supplementary Figure 3C), indicating a role in synergistically biosynthesis of leukotrienes(51). Neu.4 was characterized by *EPHB1* that regulating cell migration and adhesion(52), and *FGGY* that involved in energy metabolism (Figure 2B). Enrichment analysis disclosed that Neu.4 was involved in regulation of cell adhesion and fatty acid metabolic process (Supplementary Figure 3B). Neu.5 highly expressed G-protein beta-subunit *GNG7* and membrane repair gene *DYSF*, and genes involved in endoplasmic reticulum function and DNA damage repair like *HSPH1* (log2FC = 2.96, Pct.1 = 0.98, Pct.2 = 0.50) and *DNAJB1* (log2FC = 2.91, Pct.1 = 0.98, Pct.2 = 0.61), implying a subset of stressed neutrophil (Supplementary Figure 3B). The proportion of Neu.3 in unruptured IA tissue was particularly high (91.8%) (Figure 2C). When IA ruptured, neutrophils significantly up-regulated *S100A8/9* that involved in the regulation of cell circle and inflammation, and down-regulated *CXCL8* and *NFKBIA* and genes involved in stress mitigation (*HSPA1A, DNAJB1*) (Figure 2E), suggesting a switch in cell states.

The infiltration of monocytes/macrophages occurs at an early stage of IA, and is of pathological significance(1, 53). Monocytes/macrophages highly expressed *CD14, FCGR3A*, and type II MHC molecules and were defined as two types of monocytes (Mono.1/2), two types of macrophages (Macro.1/2), and dendritic cells (DC). These cells were enriched in pathways including phagocytosis, lysosomal activation, and antigen presentation (Figure 2D). Mono.1 expressed both *CD14* and *FCGR3A*, which may represent intermediate or non-classical monocytes, and had enriched pathways including antigen presentation, response to lipid, and positive regulation of immune response (Figure 2B, Supplementary Figure 3D). Mono.2 down-regulated *FCGR3A* and had active inflammation response and cytokine production pathways (Figure 2B, Supplementary Figure 3D), thus may be classical monocytes. Mac.1 and Mac.2 highly expressed *CD163*, with the former also expressed *CXCL3* and the latter *CD206* (Figure 2B). Although both were involved in inflammation, Mac.1 was active in response to lipopolysaccharide and cytokine stimulus, Mac.2 was enriched in endocytosis and antigen presentation (Supplementary Figure 3D). DC up-regulated *FCN1* and *EGER*, and had enriched dendrite morphogenesis pathway (Figure 2B, Supplementary Figure 3D). In the simulated evolutionary trajectory, there was significant upregulation of *CD163, MSR1* and *TNF*, and downregulation of HLA molecules from monocytes to Mac.1 and Mac.2. Given the functional plasticity of macrophages, we further interrogated the functional status of Mac.1 and Mac.2. As a result, Mac.1 and Mac.2 both performed well in predicting classical (IFN-γ-induced) and alternative activation (IL-4-induced) gene signatures and expressed similar levels of M1/M2 genes (Supplementary Figure 3E, F). In addition, monocyte/macrophage/DC had generally increased ssGSEA score of bioactive lipids-related gene signatures (Supplementary Figure 3G), which was associated with alternative activation(27). Together, these results indicated that the functional status of monocyte/macrophages under real condition was beyond M1 or M2 dichotomy.

A small amount of mast cells was identified, in line with the findings in murine aneurysm model(22). Mast cells in IA tissue highly expressed *TPSAB1, TPSB2*, and *CPA3* that encoding tryptase and metalloprotease (Figure 2B). Enrichment analysis disclosed that mast cells promoted cell proliferation and activation and had high protein kinase and lipid metabolism activity (Supplementary Figure 3H). Besides, mast cells were involved in eicosanoid metabolism, and associated with platelet aggregation, indicating a pivotal role in local inflammation.

### Lymphocytes in the IA tissue

Lymphocytes are present in normal and aneurysmal vessel walls, but their role remain less understood(20, 21, 24, 39, 54). Herein, the lymphocytes were defined as T cells (*CD3E, CD3G*), B cells (*CD79A, MS4A1*), and NK cells (*KLRF1, KLRD1*). T cell compartment was further divided into subtypes at higher resolution (Figure 3A, Supplementary Figure 4A). CD8 T cells included cytotoxic T lymphocytes (CD8 ct.1/2/3) and memory CD8 T cells (CD8 m) (Figure 3B). CD8 ct.1/2/3 highly expressed *CD8A* and *CD8B*, as well as genes involved in immune stimulation (*IFNG*) and cytotoxicity (*GZMA, GZMB*). CD8 m highly expressed *CD8B* and navïe marker *CCR7* and *SELL*(55). Genes up-regulated in CD8 T cells were involved in IL12 and TNF signaling pathway, T cell activation, and cytolysis (Supplementary Figure 4B). CD4 T cells were those highly expressed CD4 and were further defined as effector memory CD4 (CD4 em.1/2), central memory CD4 (CD4 cm), CD4 αβ, Th17 and regulatory T cells (Treg) (Figure 3B). CD4 T em.1/2 cells were characterized by lower expression of both *CCR7* and *SELL*, and higher expression of *STAT1* and *CD44*, respectively. CD4 αβ highly expressed *CCR7* and *SELL*(56). Th17 highly expressed *CD44, CCR6*, and *STAT3*, and T cell exhaustion marker *PDCD1*(56). Treg cells were characterized by *FOXP3* and *IL2RA*. CD4 T cells were involved in positive regulation of IL12 (Supplementary Figure 4E), which promotes pro-inflammatory Th1 response and the production of IFNγ by CD8 T cells(57). Similarly, Tregs had activated IL-2/STAT5 signaling pathway (Supplementary Figure 4F), which play a role in maintaining cell survival(58). Besides, mucosal associated invariant T (MAIT) cells highly expressed *RORA* and *RORC*(55) and had activated pathways including chemokine signaling and regulation of T cell differentiation (Figure 3B, Supplementary Figure 4D). Double negative T cells (T DN) expressed *CD69, CCR7* and *SELL* (Figure 3B), thus may represent a subset of central memory T cell. In addition, the unconventional T cells (T unconv) highly expressed both CD4 and CD8 molecules, and were involved in inflammation through activating IFNG and TNFA signaling pathways (Figure 3B, Supplementary Figure 4C).

**Figure 3.**
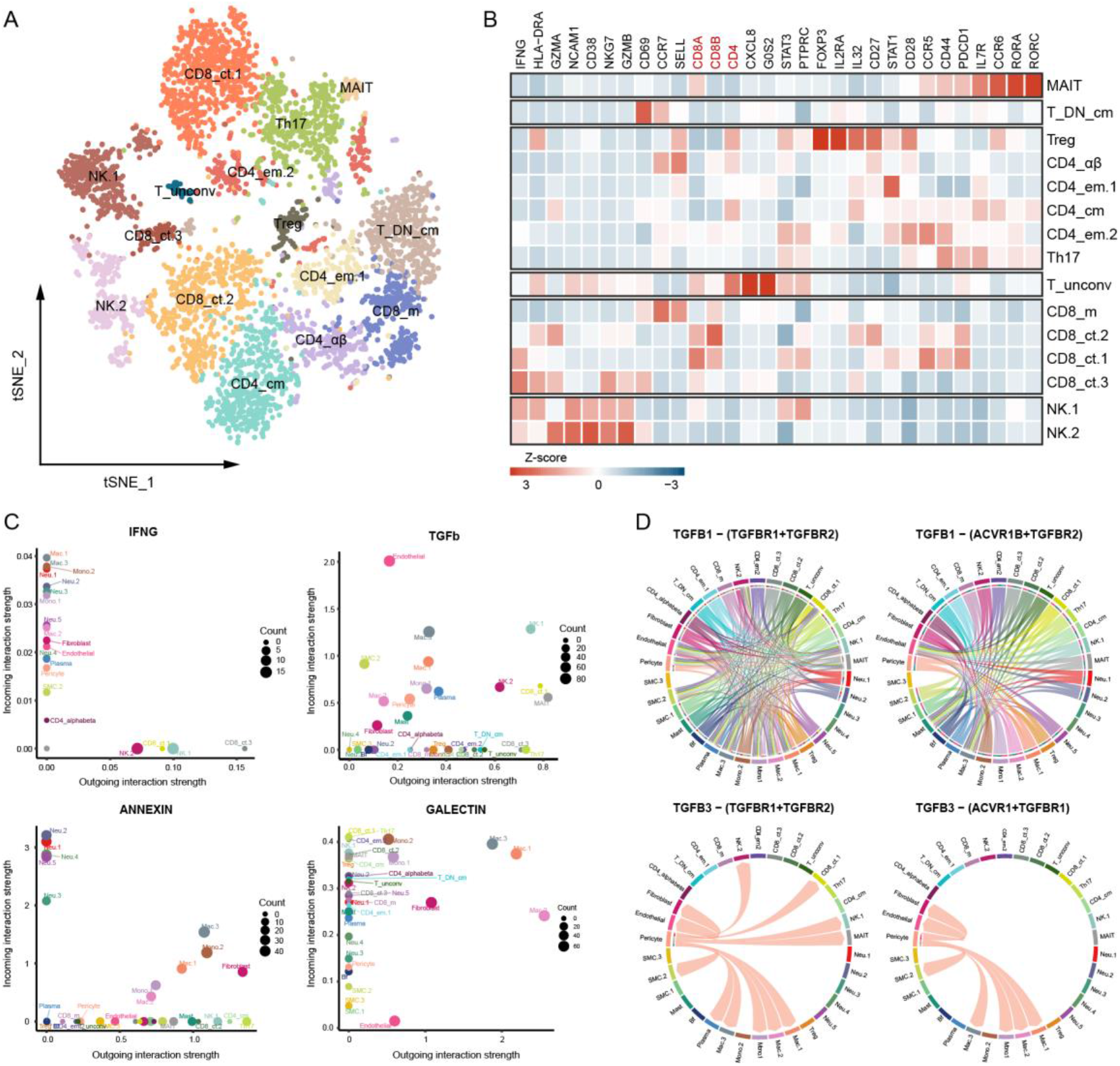
Subtypes of T lymphocytes and NK cells. (**A**) tSNE visualization of T and NK compartment. (**B**) The expression of feature genes. (**C**) Top signaling pathways mediating the interaction of T lymphocytes with other cells, including outgoing signaling IFNG: IFNG-IFNGR1/2, TGFb: TGFB1/2/3-TGFBR1/TGFBR2/ACVR1/ACVR1B, ANNEXIN: Annexin A1-FPR1/2, and incoming signaling GALECTIN: LGALS9-CD44/CD45/HAVCR2. (**D**) Details of TGFb-mediated cell-cell communications.

B cells were defined as plasma cell and Bf. Plasma cell highly expressed *CD79A* and *MS4A1* (Figure 2B), and may play a role in regulating lymphocyte activation and antigen presentation (Supplementary Figure 4G). Bf highly expressed *TNFRSF17* and *CD38* (log2FC = 0.86, Pct.1 = 0.50) and had enriched pathways including protein processing in endoplasmic reticulum and cell response to unfolded protein (Supplementary Figure 4H), indicating a stressed status. A small amount of NK cells was characterized by *KLRF1, KLRD1*, and *XCL2* and can be further divided into NK.1 and NK.2 (Figure 2B, Figure 3B). NK.1 had enriched NK cell-mediated cytotoxicity and cytolysis activity (Supplementary Figure 4I). NK.2 highly expressed *ITGAM*, encoding an integrin that mediates cell adhesion, and *NFIL3* (Supplementary Figure 4J, K), which plays a role in NK cell development(59).

To gain another glimpse of T lymphocyte function, intercellular communications were explored. The outgoing signals of T lymphocytes were prominently dependent on ANNEXIN and TGFb (Figure 3C, Supplementary Figure 5A, C). The binding of T cell ANNEXIN A1 to neutrophil FPR1/2 may be relevant to the maintenance of the latter function(60, 61). While previous studies suggested that endothelium-mediated angiogenesis was driven by pericyte-derived TGFb1(62), we found that TGFb1 was expressed by most of the T lymphocytes. Comparatively, TGFb3 signaling was restricted to the interaction of pericytes with other cells, including endothelial (Figure 3C, D). Consistent with previous reports, IFNG was primarily produced by CD8 T cells and NK cells and acted on macrophages and neutrophils. The binding of LGALS9 (Galectin) to corresponding receptors HAVCR2/CD44/CD45 may be relevant to avoid overactivation of cytotoxic T lymphocytes (Figure 3C, Supplementary Figure 5B) (63, 64).

### Inflammatory features associated with IA rupture

To elucidate inflammatory features associated with IA rupture, two external datasets were retrieved. The ssGSEA scores of Mac.1, Mac.2, DC, and Mono.2 were significantly increased in the ruptured IA tissue (Figure 4A, B, Supplementary Table 4), and the ssGSEA score of Mac.1 gave the best performance in predicting IA rupture (Figure 4C, D), in accordance with previous findings that specific type of monocyte/macrophage was implicated in bleeding of cerebral arteriovenous malformations and thoracic aortic aneurysm(20, 23). Given the plasticity of macrophage phenotype, further analysis was conducted to address the transcriptional features of Mac.1. As a result, Mono.1 and Mac.1 shared similar transcriptional regulatory patterns (Figure 4E, Supplementary Figure 6A). NF-κB (*NFKB1, NFKB2* and *RELB*) was activated in Mac.1, together with its negative regulator *HIVEP2*(65) (Figure 4F). The transcriptional networks of Mac.1 and Mac.2 were distinct, and the high-confidence target genes of Mac.1 were enriched in TNF signaling pathway, unlike that of Mac.2(Supplementary Figure 6B-D). Moreover, WGCNA algorithm identified the intrinsic gene expression pattern of Mac.1, some of which were involved in aneurysm progression and rupture (Supplementary Figure 7). Together, these results suggested that Mac.1 was involved in IA rupture.

**Figure 4.**
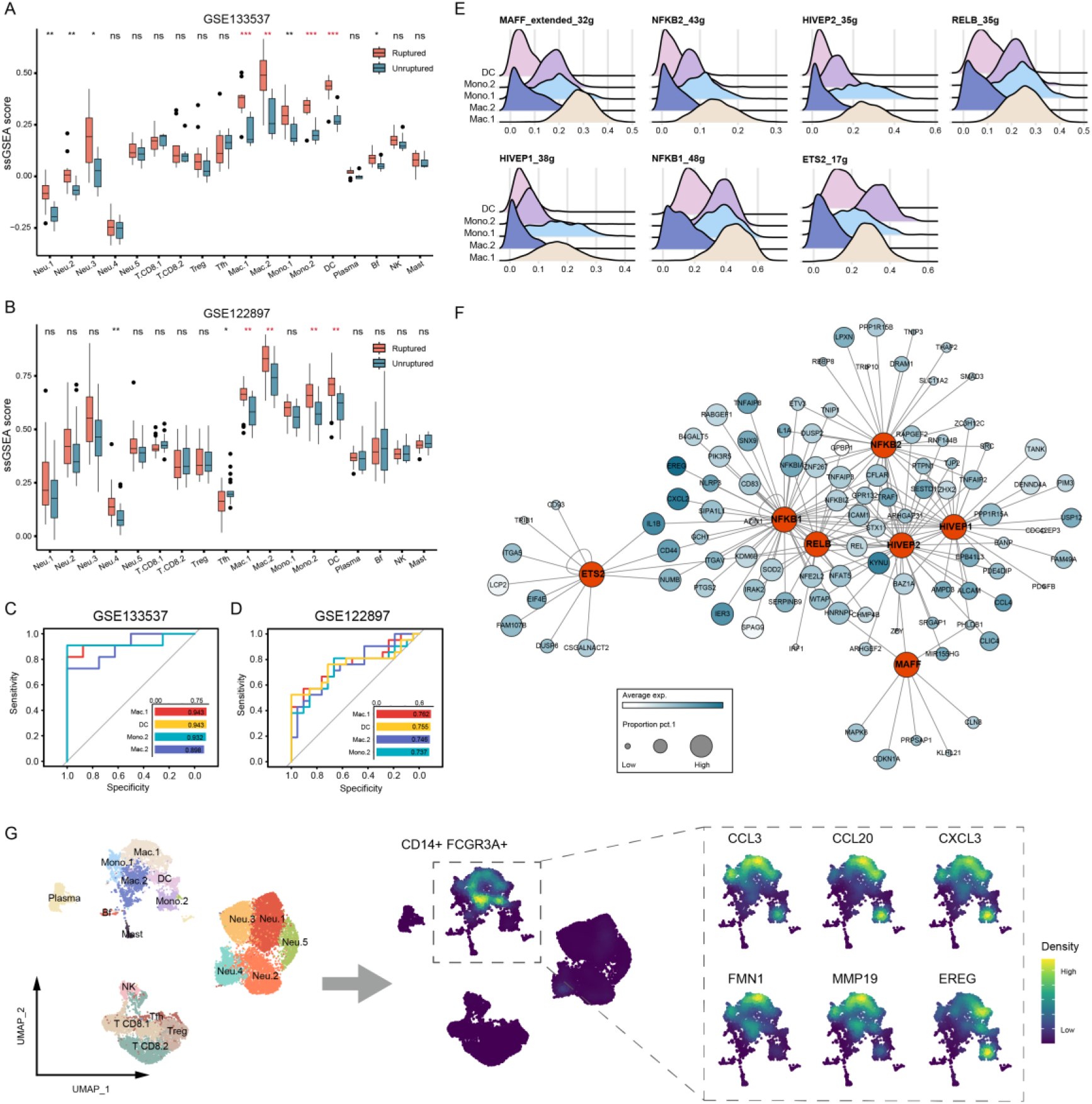
Macrophages implicated in IA rupture. (**A, B**) ssGSEA score of immune cells in ruptured and unruptured IA tissues. Genes with log2FC > 0.5 and Pct.2 < 0.15 in each cell type are used as signature genes. (**C, D**) Performance of gene signatures in predicting IA rupture. (**E**) The activated transcriptional factors in monocytes/macrophages. (**F**) The network of transcriptional factors mainly activated in Mac.1 and their high-confidence target genes. (**G**) Density plot showing feature genes of Mac.1.

To interrogate how Mac.1 may facilitate IA rupture, the intercellular communications mediated by Mac.1 were estimated using cellphoneDB. The interactions between Mac.1 with vascular cells (including endothelial, pericyte, SMC, and fibroblast) were largely depend on TNFα (encoded by *TNF*) (Figure 5A, B). TNFα is functionally pleiotropic depending on the receptors. For instance, binding to TNFR2 (encoded by *TNFRSF1B*) maintain cell survival, while binding to TNFR1 (encoded by *TNFRSF1A*) or FAS induce apoptosis(66). Although *FAS* and *TNFRSF1A* were expressed by vascular cells (SMC.2, endothelial, pericyte, and fibroblast) and some immune cells, TNFα-mediated cell death may be limited to vascular cells due to the higher expression of *TNFRSF1B* in immune cells (Figure 5C). Previous hypothesis suggests that the aneurysmal wall inflammation exists prior to IA rupture (3, 10). We found that macrophages characterized by *NFKB1* and *HIVEP2* transcriptional activation were also present in the unruptured sample (Mac.1/2) (Figure 5D-F, Supplementary Figure 8A). These cells were significantly enriched in the TNF signaling pathway and interacted with SMCs through the binding of TNF and TNFSF13 to FAS and TNFRSF1A (Figure 5G, Supplementary Figure 8B).

**Figure 5.**
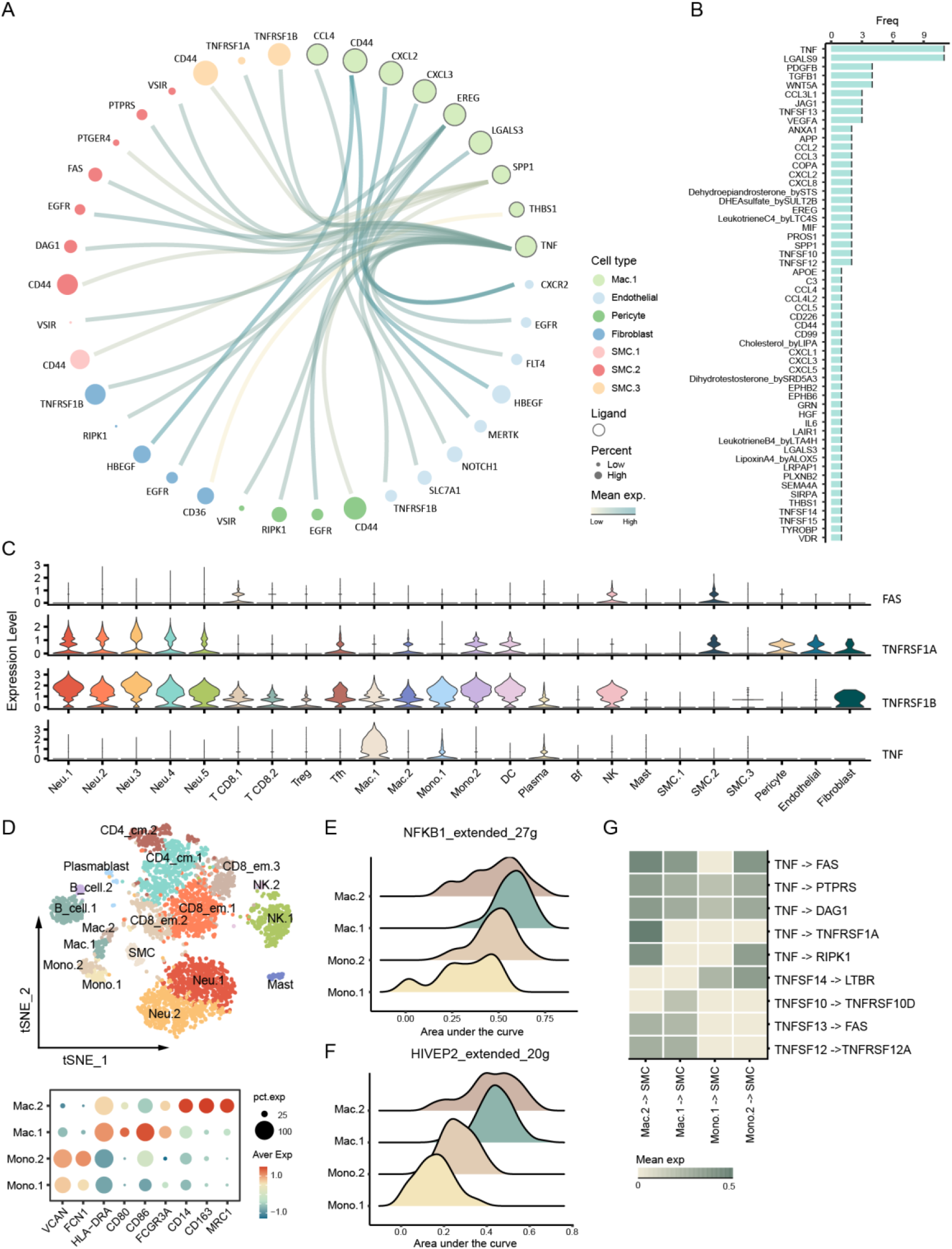
Interactions between Mac.1 and vascular cells. (**A**) Circus plot exhibiting top expressed ligands of Mac.1 and their corresponding receptors of vascular cells. Bubble size is proportional to the percentage of cells expressing corresponding gene (Pct.1). The color of line is proportional to the mean expression of ligand-receptor pairs estimated by cellphoneDB. (**B**) Frequency of ligands of Mac.1 that interact with vascular cells. (**C**) Expression of ligands and receptors associated with TNFα-mediated cell survival and death. (**D**) tSNE visualization of cells in the unruptured IA tissue and their feature genes. (**E, F**) The activation of NFKB1 and HIVEP2 in Mac.1/2 of the unruptured IA. (**G**) The TNF family-mediated interactions between Mac.1/Mac.2/Mono.1/Mono.2 and SMCs in unruptured IA.

Despite the involvement of multiple immune cells and inflammatory mediators in IA wall inflammation, Mac.1 expressed the highest amounts of ligands (outgoing signaling strength) and receptors (incoming signaling strength) (Figure 6A), thus serving as a hub of the inflammation network. We identified the top ligands expressed by Mac.1 and their corresponding receptors, revealing that the prominent interaction patterns of Mac.1 involved low expression of *TNF* to bound to *TNFRSF1A/B, FAS*, and *ICOS* (Figure 6B, C), a co-stimulatory receptor for T lymphocytes(67), and high expression of various chemokines, such as *CXCL8* which recruit neutrophils and contribute to the destruction of IA tissue(68). In terms of macrophage recruitment, CCL3, a known macrophage chemotactic factor, was mainly derived from monocytes/macrophages and inflammatory SMC (SMC.3) (Figure 6D). In addition, CCL4(69), CCL5(70), MIF(71, 72), as well as CCL3L1 and CCL4L2 that can bind to CCR5(73, 74), were all involved in macrophage recruitment. Even in the unruptured sample, Mac.1/2 maintained the highest intensity of intercellular communication (Figure 6E). Mac.1/2 were involved in inflammation in a TNF and chemokine-dependent mechanism (Figure 6F, G), and may be recruited by monocytes/macrophages, lymphocytes, and smooth muscle cells by producing similar chemokines (Supplementary Figure 8F, G). Together, these results highlighted that macrophage-mediated inflammation preceded IA rupture.

**Figure 6.**
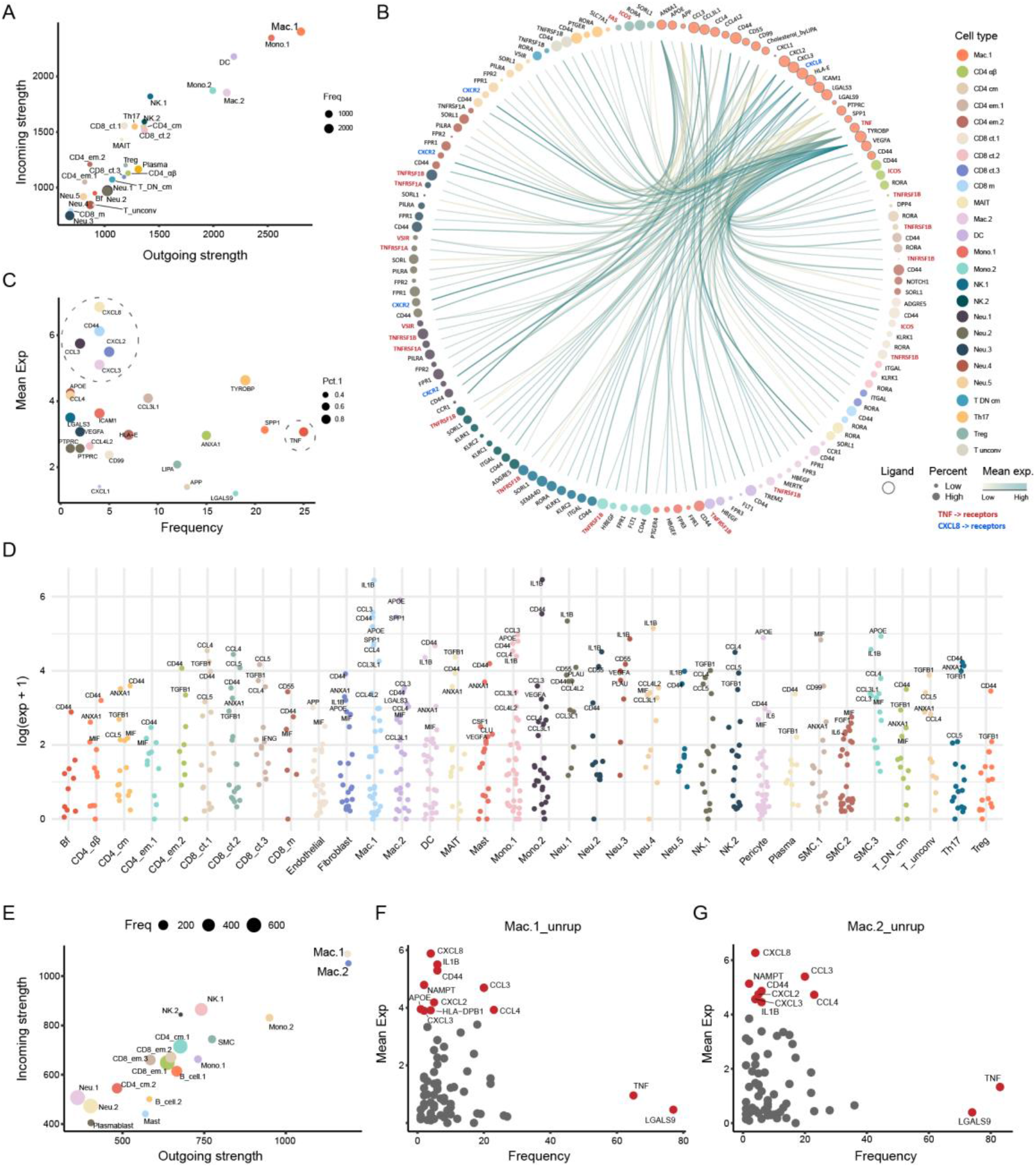
Interactions between Mac.1 and other immune cells. (**A**) The interaction strength of each type of cell. Incoming strength, count of statistically significant receptors. Outgoing strength, count of statistically significant ligands. (**B**) Top 25 ligand-receptor pairs mediating interactions between Mac.1 and other immune cells. Bubble size is proportional to the percentage of cells expressing corresponding gene (Pct.1). The color of line is proportional to the mean expression of ligand-receptor pairs estimated by cellphoneDB. TNF and CXCL8-mediated interactions are annotated in red and blue, respectively. (**C**) The strength of ligands of Mac.1. Frequency represents the count of statistically significant ligands among the top 25 ligands-mediated cell-cell interactions. Genes with high frequency of intercellular communication or high expression were circled. (**D**) Ligands expressed by other immune cells that bind to receptors of Mac.1. (**E**) The interaction strength of each type of cell in unruptured IA. (**F, G**) The strength of Mac.1/2 expressed ligands of the unruptured IA. Frequency represents the count of statistically significant ligands overall. Unrup, unruptured.

## Discussion

IA may represent a heterogeneous group of diseases manifested by pathological dilation of intracranial arteries, in which the inflammation plays an essential role in disease progression. This study provided a detailed cell atlas of human IA tissue and highlights the inflammation features implicated in rupture. We expanded cellular diversity of human IA through the identification of pericyte, and provided a compendium of immune cell types along with functional characteristics. Notably, we identified a subset of macrophage that was closely associated with IA rupture, with inflammatory activity being its major functional orientation. The macrophage-mediated inflammatory response patterns coexisted in ruptured and unruptured IA and are therefore promising therapeutic targets for unruptured IA.

Recent studies thoroughly characterized the major cell types and corresponding transcriptome features of the cerebrovascular system, and identified endothelial, smooth muscle cell, pericyte, perivascular fibroblast, and less common fibromyoblast(20, 21). These major cell types also exist in human IA tissue(24). Interestingly, we identified pericytes in the IA wall for the first time, which previous studies have shown to exist in small vessels or capillaries(75, 76). Pericyte is morphologically divided into reticular and fascicular subtypes, and play a role in regulating blood flow and maintaining the brain-blood barrier(75, 77). Endothelin and PDGFBR expressed by endothelial cells recruit bone marrow-derived pericytes, which, subsequently promote endothelial cell differentiation and neovascular sprouting via the TGFβ pathway(45, 62). Consistent with these findings, we found that pericytes engaged in a TGFB1/3-dependent interaction with endothelial cells, where TGFB1 is predominantly derived from T lymphocytes and TGFB3 exclusively from pericytes. Recently, vasa vasorum has been identified in human IA tissue, which coincides with the scenario of endothelial-pericyte interaction-mediated angiogenesis(44, 45, 62, 78). These were interesting findings for understanding the pathology of the IA wall, and the presence of neovascularization and the role of pericytes in maintaining vascular stability raises the possibility of an association between bleeding within the IA wall and pericytes dysregulation.

Astounding number of immune cells were identified in both unruptured and ruptured IA tissues, consistent with previous findings(7, 9, 79). Immune cells including T and B lymphocytes, monocyte/macrophage, neutrophil, and to a lesser extent NK cell and mast cell are involved in IA rupture(22, 24, 68, 79). Particularly, monocyte/macrophages can be found at an early stage of the disease(7, 22, 24), and according to our results, lymphocytes and inflammatory SMC were involved in monocyte/macrophages recruitment. There presented a subset of macrophages that was characterized by the production of TNFα and chemokines that recruit neutrophils, both serving as catalysts for IA rupture(68, 80). Using multiple algorithms, we found that macrophages associated with IA rupture had NF-κB activation and express a range of marker genes. TNFα is a well-known activator of macrophage NF-κB, and can be initially secreted by lymphocytes(80, 81). The activation of NF-κB enables macrophages to upregulate various downstream immune-related genes, thereby amplifying the inflammatory response. Therefore, it appears that NF-κB activated macrophages hold promise as therapeutic targets. However, macrophages can be recruited, activated by chemokines and cytokines of multiple cellular origin, making a better understanding of the systemic characteristics of the inflammatory response in IA wall a prerequisite for designing rational targets.

In summary, we compiled a catalogue of cells and their gene expression features of human IA wall and identified a specific type of macrophage as potential therapeutic for unruptured IA. Due to the importance of inflammation and current paucity in understanding of the impact of immune cells on IA, further studies should be dedicated to elucidating how the inflammation in the IA wall is orchestrated.

## Data Availability

The source code and collated gene expression profile can be obtained from the corresponding author upon reasonable request.

## Abbreviations

CCA: canonical correlation analysis
DC: dendritic cell
IA: intracranial aneurysm
MAIT: mucosal associated invariant T
MNN: mutual nearest neighbor
PCA: principal component analysis
SMC: smooth muscle cell
SAH: spontaneous subarachnoid hemorrhage
ssGSEA: single-sample gene set enrichment analysis
WGCNA: weighted gene co-expression network analysis.

## Acknowledgements

The authors gratefully thank the patients participated this program, and prof. Wang Yuan for his support in data analysis.

## Author contribution

Study design: Yi Liu, Hang Ji, and Yue Li. Data collection and curation: Yue Li, Haogeng Sun, Ruiqi Chen, and Anqi Xiao. Data analysis: Hang Ji, Ran Zhou, and Yue Li. Manuscript drafting: Hang Ji and Yue Li. Manuscript revise: Rong Wang, Yongbo Yang, and Chao You. Study supervising: Yi Liu and Chao You. Hang Ji and Yue Li contributed equally to this study.

